# Validating risk prediction models for multiple primaries and competing cancer outcomes in families with Li-Fraumeni syndrome using clinically ascertained data at a single institute

**DOI:** 10.1101/2023.08.31.23294849

**Authors:** Nam H. Nguyen, Elissa B. Dodd-Eaton, Jessica L. Corredor, Jacynda Woodman-Ross, Sierra Green, Nathaniel D. Hernandez, Angelica M. Gutierrez Barrera, Banu K. Arun, Wenyi Wang

**Affiliations:** The University of Texas MD Anderson Cancer Center, Department of Bioinformatics and Computation Biology, Houston, TX; Rice University, Department of Statistics, Houston, TX; The University of Texas MD Anderson Cancer Center, Department of Clinical Cancer Genetics, Houston, TX; The University of Texas MD Anderson Cancer Center, Department of Breast Medical Oncology, Houston, TX

## Abstract

**Purpose:** There exists a barrier between developing and disseminating risk prediction models in clinical settings. We hypothesize this barrier may be lifted by demonstrating the utility of these models using incomplete data that are collected in real clinical sessions, as compared to the commonly used research cohorts that are meticulously collected.

**Patients and methods:** Genetic counselors (GCs) collect family history when patients (i.e., probands) come to MD Anderson Cancer Center for risk assessment of Li-Fraumeni syndrome, a genetic disorder characterized by deleterious germline mutations in the *TP53* gene. Our clinical counseling-based (CCB) cohort consists of 3,297 individuals across 124 families (522 cases of single primary cancer and 125 cases of multiple primary cancers). We applied our software suite LFSPRO to make risk predictions and assessed performance in discrimination using area under the curve (AUC), and in calibration using observed/expected (O/E) ratio.

**Results:** For prediction of deleterious *TP53* mutations, we achieved an AUC of 0.81 (95% CI, 0.70 – 0.91) and an O/E ratio of 0.96 (95% CI, 0.70 – 1.21). Using the LFSPRO.MPC model to predict the onset of the second cancer, we obtained an AUC of 0.70 (95% CI, 0.58 – 0.82). Using the LFSPRO.CS model to predict the onset of different cancer types as the first primary, we achieved AUCs between 0.70 and 0.83 for sarcoma, breast cancer, or other cancers combined.

**Conclusion:** We describe a study that fills in the critical gap in knowledge for the utility of risk prediction models. Using a CCB cohort, our previously validated models have demonstrated good performance and outperformed the standard clinical criteria. Our study suggests better risk counseling may be achieved by GCs using these already-developed mathematical models.

## Introduction

Li-Fraumeni syndrome (LFS) is a hereditary cancer syndrome identified by deleterious germline mutations in the *TP53* tumor suppressor gene^1^. Patients with LFS are at significantly increased risks of a spectrum of cancer types, most notably early-onset breast cancer, soft-tissue sarcoma, osteosarcoma, among many others^1–3^. The lifetime risks of cancer are 93% and 73% for women and men respectively^4^, with a 50% risk of second primary cancer for individuals that have been diagnosed with a first primary^5^. Conversations with these patients regarding important decisions such as genetic testing and cancer screening have been challenging, partly because genetic counselors (GCs) could only provide general, as compared to personalized, cancer risks associated with LFS^6^. Although risk prediction models have been developed for other hereditary cancer syndromes, such as BRCA1/2 and breast cancer^7–9^, LFS remained an untouched area until recently. To facilitate personalized risk predictions in clinics, we developed two models specifically for families with LFS: (i) a competing-risk model that predicts cancer-specific (CS) risks for the first primary cancer^10^, and (ii) a recurrent event model that extends the risk prediction to multiple primary cancer (MPC)^11^. These models were trained on an LFS cohort rich in family history, and successfully validated on independent cohorts^12,13^.

The datasets used to train and validate these risk prediction models were research protocol-based (RPB). RPB refers to data that are collected via rigorous procedures to obtain complete and accurate patient cohorts for research purposes (**Figure 1**). The study investigators contact eligible patients for data collection via extensive use of questionnaires and phone interviews. Additional follow-ups are conducted on a regular basis to fill in the missing data and to acquire new incidences of cancer diagnoses of the participant or family members, the latest births or deaths within the family, and any additional germline testing information. This is a diligent data collection process that could take over 20 to 30 years^14–16^. For this reason, research datasets are ideal for training statistical models to estimate key epidemiological parameters of a study population.

**Figure 1:**
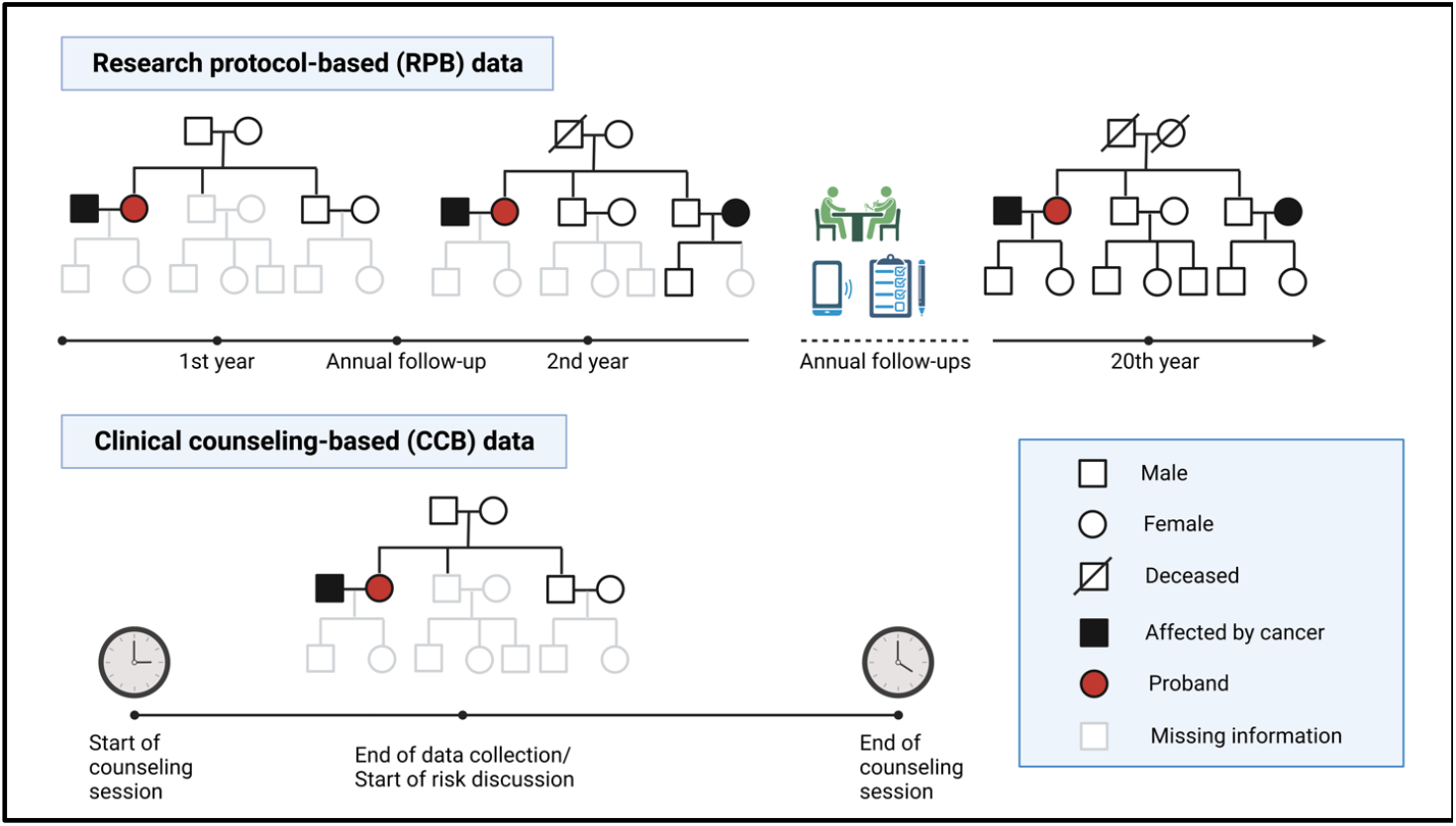
Comparison of the data collection process for RPB and CCB cohorts. RPB data are collected and updated over an extended period of time to ensure completeness and accuracy for research purposes, whereas CCB data represent a snapshot of information taken by genetic counselors over ∼20 minutes during 1-hour counseling sessions.

RPB data, however, do not represent datasets that are typically observed and collected in clinical settings, where patients with cancer history that is indicative of LFS make appointments with medical professionals for in-depth risk assessment. Given this scenario, we use the term clinical counseling-based (CCB) to refer to the data that are routinely collected by GCs during counseling sessions (**Figure 1**). CCB differs significantly from RPB because patients may not have accurate and complete family histories and some families have younger members who have not developed cancer. This leads to a higher rate of missing information in CCB datasets such as previous genetic testing results, ages of death, and ages at cancer diagnoses. Based on this snapshot of family history (collected over ∼20 minutes), GCs perform a comprehensive risk assessment, communicate these risks with the patients, and potentially recommend at-risk individuals for further testing and screening.

Due to the wide discrepancies in data quality between the CCB and RPB datasets, it is important to determine whether statistical models that are trained and validated on RPB cohorts can perform well enough on a CCB cohort to be useful in real clinical settings. Given the large number of risk prediction models for hereditary cancer syndromes^7–9^, it is surprising to see very few that made their way into the clinics^17^. One potential reason is these models were mostly validated using well established research databases or registry data^18–23^ rather than clinical data^24^. Thus, a successful validation using CCB cohorts may represent a key ingredient to break the barrier to clinical utility. In this paper, we perform a clinical validation study of our risk prediction models on a CCB cohort of 124 families who underwent genetic counseling at the Clinical Cancer Genetics (CCG) program at MD Anderson Cancer Center (MDACC) between 2000-2020.

## Materials and methods

### Patient cohorts

Using a collection of 189 families that were recruited through probands diagnosed with pediatric sarcoma at MDACC from 1944 to 1982^14–16^, we have estimated the model parameters for risk prediction^10,11^. We refer the readers to **Data Supplement** for detailed descriptions of this training dataset.

The validation dataset were collected on *TP53* mutation carriers from CCG program at MDACC. Personal and family history were collected during a genetic counseling session and immediately entered into the patient’s electronic medical record. Data were automatically pulled into a Progeny database used by the CCG program for tracking families. This database includes patients counseled starting from year 2000 to 2020. For this study, only patients that were identified to have a pathogenic or likely pathogenic germline mutation in *TP53* through single-gene testing or multi-gene panel were included. Patients that did not meet the Classic^3^ or Chompret^25,26^ LFS criteria were tested either because of clinical suspicion from a certified GC or they were identified on panel testing performed on suspicion for other hereditary cancer syndromes. Testing was performed in several CLIA/CAP certified laboratories. Family members of the confirmed *TP53* mutation carrier were not required to undergo additional testing, however recommendations for family member testing were made during standard of care genetic counseling sessions. This cohort includes a total of 124 families and 3,297 family members. Summaries of both datasets are given in **Table 1**.

**Table 1:** Categorization of all family members in the research cohort (189 families) used as training data and the clinical cohort (124 families) used as validation data by gender, number of primary cancers and mutation status. SPC = single primary cancer, MPC = multiple primary cancer, WT = wildtype, Mut = TP53 mutation, U = unknown

|  | Research cohort (training data) |  |  |  | Clinical cohort (validation data) |  |  |  |
| --- | --- | --- | --- | --- | --- | --- | --- | --- |
|  | WT | Mut | U | Total | WT | Mut | U | Total |
| **Male** |  |  |  |  |  |  |  |  |
| Healthy | 295 | 9 | 1,276 | 1,580 | 17 | 13 | 1,376 | 1,406 |
| SPC | 105 | 25 | 139 | 269 | 3 | 15 | 210 | 228 |
| MPC | 3 | 14 | 8 | 25 | 1 | 10 | 20 | 31 |
| Subtotal | 403 | 48 | 1,423 | 1,874 | 21 | 38 | 1,606 | 1,665 |
| **Female** |  |  |  |  |  |  |  |  |
| Healthy | 341 | 8 | 1,207 | 1,546 | 21 | 20 | 1,203 | 1,244 |
| SPC | 120 | 21 | 102 | 243 | 3 | 33 | 260 | 296 |
| MPC | 4 | 19 | 10 | 33 | 1 | 59 | 32 | 92 |
| Subtotal | 465 | 48 | 1,319 | 1,832 | 25 | 112 | 1,495 | 1,632 |
| **Total** | 868 | 96 | 2,742 | 3,706 | 46 | 150 | 3,101 | 3,297 |

### Risk prediction models

We previously developed and validated two models for LFS risk predictions^10–13^. The CS model^10^ estimates the cancer-specific age-at-onset penetrance, defined as the probability of developing a particular cancer type prior to all others by a certain age given the patient’s covariates and cancer history. Let *K* be the number of cancer types. We model the hazard function of each cancer type via frailty modeling^27^ as follows

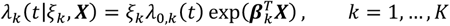

where *λ*_0,*k*_(*t*) is the baseline hazard, and the frailty term, *ξ*_*k*_, is modeled via *Gamma*(*ν*_*k*_, *ν*_*k*_). We allow the hazard function to depend on patient-specific covariates ***X*** = {*G, S, G* × *S*}^T^, where *G* denotes the *TP53* mutation status (1 for mutation and 0 for wildtype) and *S* denotes the gender (1 for male and 2 for female). Under this modeling framework, we compute the family-wise likelihood using the peeling algorithm^28^, followed by ascertainment bias correction^29^, and finally estimate the regression coefficients ***β***_*k*_ via Markov chain Monte Carlo. The age-at-onset penetrance for the *k*-th cancer type is then given by

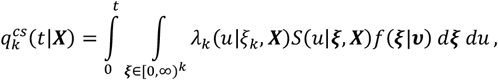

where *f* is a Gamma density function, and 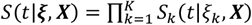 with 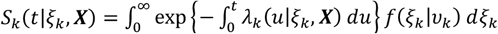. In our LFS application, we consider three competing cancer types: (1) sarcoma, including soft-tissue and osteosarcoma (*k* = 1), (2) breast cancer (*k* = 2), and (3) all other cancer types combined (*k* = 3). We also include death (*k* = 4) as another competing risk.

We further developed the MPC model^11^ to estimate the MPC-specific age-at-onset penetrance, defined as the probability of developing the next primary cancer by a certain age given the patient’s covariates and cancer history. We model the occurrence process of cancer using a non-homogenous Poisson process to capture the age-dependency of cancer risks over a patient’s lifetime^30,31^. Let *L* be the number of primary cancers. We model the intensity function via frailty modeling as before

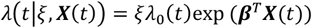

where *λ*_0_(*t*) is the baseline intensity, and *ξ* is the frailty term that is modeled via *Gamma*(*ν, ν*). We use patient-specific covariates ***X***(*t*) = {*G, S, G* × *S, D*(*t*), *G* × *D*(*t*)}^T^, where we introduce *D*(*t*), an indicator variable that indicates whether a patient has developed a primary cancer before time *t*, to allow the risks of subsequent primary cancers to depend on the first^32–34^. Let *T*_*l*_ be the time of the *l*-th cancer occurrence, *l* = 1, …, *L*. Given the estimated model parameters, the age-at-onset penetrance for the *l*-th primary cancer is then given by

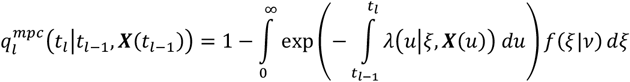

where *f* is a Gamma density function. We only model up to the second primary due to limited occurrences of the third primary and beyond.

Most patients do not undergo genetic testing due to the burdens of such procedure (i.e., *G* is unknown). Both models utilize the BayesMendel method^35^ to infer the probability of carrying a deleterious *TP53* variant for these patients based on their family history. Given a patient with unknown genotype *G*_0_ and history ***H*** = {*H*_1_, …, *H*_*n*_} of the *n* family members, our goal is to estimate *P*[*G*_0_|***H***]. Following our previous study^36^, we set the prevalence of pathogenic *TP53* mutations in the general population, denoted by *P*[*G*_0_ = 1], to be 0.0006. Assuming the Hardy-Weinberg equilibrium, it follows that the prevalence of wildtype (*G*_0_ = 0), heterozygous mutation (*G*_0_ = 1) and homozygous mutation (*G*_0_ = 2) are 0.9988, 0.0005996 and 3.6e-07, respectively. We provide the detailed computations of *P*[*G*_0_ = *g*|***H***], *g* ∈ {0,1,2}, in **Data Supplement**. The cancer-specific risks of this patient are given by weighted sums of the corresponding penetrance estimates

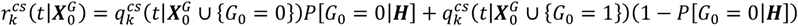

where 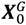 denotes the covariates without genotype. The MPC risk, denoted by 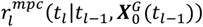, can be computed from the MPC-specific penetrance estimates in a similar way.

### Validation Study Design

We excluded family members who had either (i) unknown ages at cancer diagnoses for the first or second primary cancer, or (ii) unknown ages at last contact if they had never had cancer, or both, from the set of validation subjects. Missing information among the excluded family members can still negatively impact performance on the validation subjects because the key assumption of our models lies in the Mendelian inheritance pattern that is implicitly demonstrated by cancer outcomes within the family^36^. The goal is to evaluate whether our models are sufficiently robust to incomplete datasets that arise in clinical settings.

We first validated the utility of our models to predict an individual’s probability of carrying a deleterious *TP53* mutation given the family history collected during a genetic counseling session. To do that, we used the models to make predictions for the validation subjects, including the probands, that had undergone genetic testing, then compared the predicted outcomes with the confirmed genotypes. In the calculations, we disregarded all testing results. This mimicked a real scenario, in which GCs use the models to assess the risks of the probands, and to identify at-risk individuals within their families for recommendation of screening and testing. We then conducted a similar validation, in which we made the predictions for non-proband family members given the confirmed genotypes of the probands, to evaluate the impacts of this additional information.

Next, we ran the models to make cancer risk predictions for patients in the CCG dataset. We further excluded the probands due to ascertainment bias. For the MPC model, we divided the validation subjects into three groups: those without cancer (group 1), those with SPC (group 2), and those with MPC (group 3). We then validated the model in two tasks: (i) to predict individuals with at least one primary cancer versus those without, and (ii) to predict individuals with MPC versus those with SPC. For the first task, we recorded the ages at last contact for individuals in group 1, and the ages at first cancer diagnosis for those in groups 2 and 3. For each individual, we computed the risk probability to develop a first primary cancer at the recorded age *t*_1_. By varying the cutoff on the risk estimates and comparing the predictions with the actual outcomes (at least one cancer versus no cancer), we constructed the receiver operating characteristic curve (ROC) and calculated the area under the curve (AUC). For comparison, we also used the Kaplan-Meier (KM) method to achieve the same prediction objective. For the second task, we recorded the ages at last contact for individuals in group 2, and the ages at second primary cancer diagnosis for those in group 3. We computed the risk probability to develop a second primary cancer at the recorded age *t*_1_ given the covariates and cancer history up to age *t*_2_. We constructed the ROC curve in the same way as described above.

For the CS model, we recorded the age at first event (i.e., the age at diagnosis of the first primary cancer if the individual had a cancer history, or the age at last contact if the individual had never had cancer). We used the model to compute the risk probability at the recorded age *t*_1_ for each of the four competing outcomes (i.e., sarcoma, breast cancer, all other cancer types, and mortality). We followed the same steps to construct ROC curves for predicting one cancer type versus all other outcomes.

In addition to AUCs, which provide a comprehensive measure of the model’s ability to discriminate between binary outcomes, we also assessed model calibration via the observed/expected (O/E) ratios. The 95% confidence intervals for the performance metrics were computed via bootstrapping.

## Results

### Comparison of clinical and research data

Our model training dataset, being RPB, was meticulously collected via rigorous research protocols to obtain information that is as accurate and complete as possible for research purposes. On the other hand, the CCG dataset, being CCB, represented snapshots of information taken by GCs during their counseling sessions. **Table 2** highlights the main differences, most notably the level of missing data, between RPB and CCB based on the key summary statistics of these two datasets (all comparisons presented a Chi-square test *P* < 0.001).

**Table 2:** Comparison of a research cohort (Pediatric Sarcoma as training data) and a clinical cohort (CCG as validation data) on the extent of missing ages at last contact and missing ages at cancer diagnoses at both family and individual levels. Summary statistics for the number of individuals per family are reported to contrast the depth of data collection procedures in research and clinical cohorts as they happen in the unit of families

|  | **RPB training data** | **CCB validation data** |
| --- | --- | --- |
| **Number of families** |  |  |
| **All family members** |  |  |
| Complete data | 189 (100%) | 10 (8%) |
| Missing ages at last contact only | 0 (0%) | 46 (37%) |
| Missing ages at cancer diagnosis only | 0 (0%) | 0 (0%) |
| Missing both ages at last contact and ages at cancer diagnosis | 0 (0%) | 68 (55%) |
| **Total** | **189** | **124** |
| **Chi-square test** | **P < 0.001** |  |
| **First-degree relatives and spouse only** |  |  |
| Complete data | 189 (100%) | 68 (55%) |
| Missing ages at last contact only | 0 (0%) | 41 (33%) |
| Missing ages at cancer diagnosis only | 0 (0%) | 10 (8%) |
| Missing both ages at last contact and ages at cancer diagnosis | 0 (0%) | 5 (4%) |
| **Total** | **189** | **124** |
| **Chi-square test** | **P < 0.001** |  |
| **Number of individuals** |  |  |
| **All family members** |  |  |
| Complete data | 3,706 (100%) | 1,748 (53%) |
| Missing ages at last contact only | 0 (0%) | 1,339 (41%) |
| Missing ages at cancer diagnosis only | 0 (0%) | 138 (4%) |
| Missing both ages at last contact and ages at cancer diagnosis | 0 (0%) | 72 (2%) |
| **Total** | **3,706** | **3,297** |
| **Chi-square test** | **P < 0.001** |  |
| **First-degree relatives and spouse only** |  |  |
| Complete data | 1,126 (100%) | 487 (79%) |
| Missing ages at last contact only | 0 (0%) | 105 (17%) |
| Missing ages at cancer diagnosis only | 0 (0%) | 19 (3%) |
| Missing both ages at last contact and ages at cancer diagnosis | 0 (0%) | 2 (0.3%) |
| **Total** | **1,126** | **613** |
| **Chi-square test** | **P < 0.001** |  |
| **Number of individuals per family** |  |  |
| Min | 3.00 | 1.00 |
| 5% percentile | 4.00 | 1.00 |
| 10% percentile | 4.80 | 4.00 |
| 25% percentile | 6.00 | 16.00 |
| Median | 7.00 | 26.50 |
| Mean | 19.69 | 26.59 |
| 75% percentile | 10.00 | 36.00 |
| 90% percentile | 14.80 | 48.00 |
| 95% percentile | 72.20 | 53.85 |
| Max | 719.00 | 75.00 |

### Validation of *TP53* mutation prediction

In **Figure 2a**, we compared the models’ performance in predicting the probability of *TP53* mutations with the Classic^3^ and Chompret^25,26^ criteria, which are currently recommended in the National Comprehensive Cancer Network (NCCN) guidelines (version 3.2023) for LFS. Our CS and MPC models achieved AUCs of 0.76 (95% CI, 0.68 – 0.84) and 0.78 (95% CI, 0.71 – 0.85) respectively. In particular, with a decision threshold of 0.22, the MPC model achieved a true positive rate (TPR) of 0.75 and a false positive rate (FPR) of 0.29, while the Chompret criteria achieved a near-zero FPR at the cost of a low TPR. For calibration, the MPC model achieved a much better O/E ratio of 1.66 (95% CI, 1.53 – 1.80) compared to the CS model, which showed underestimation with an O/E ratio of 7.83 (95% CI, 7.20 – 8.47). The results showed that the MPC model performed better than the CS model in both criteria, thus providing further support for selecting the MPC model as default in our clinical risk prediction tool LFSPRO^36^.

**Figure 2:**
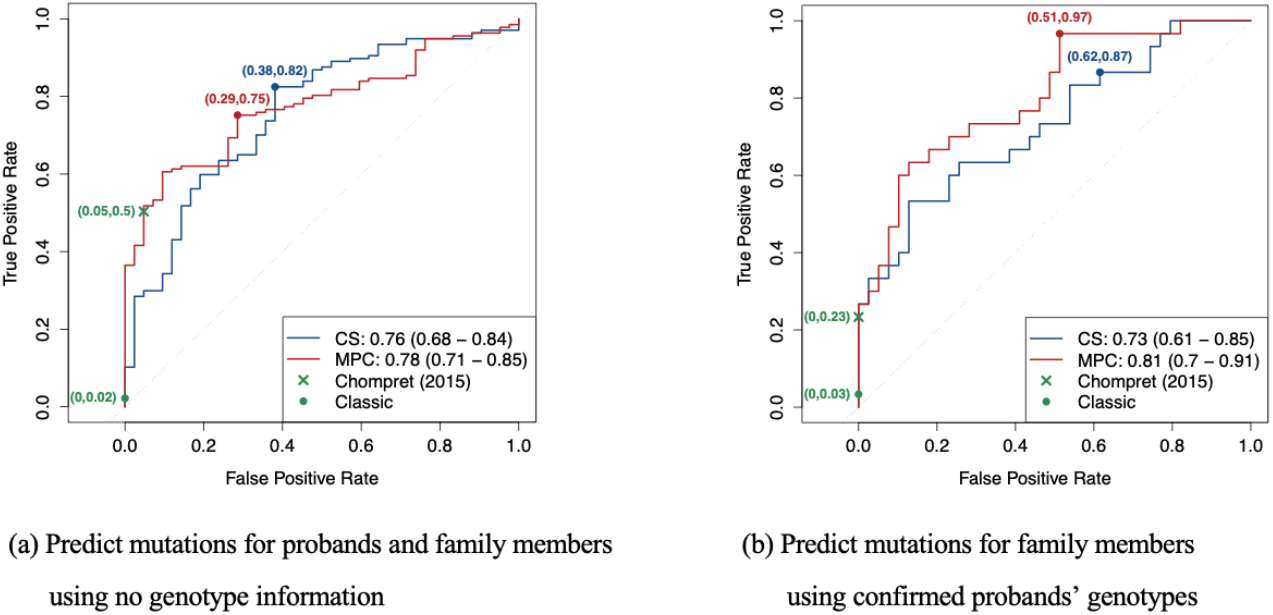
ROC curves, and the 95% bootstrapped confidence intervals of the AUCs, for TP53 mutation predictions in the CCG cohort using the CS and MPC models under two scenarios: (a) predict mutations for both the probands and their family members when no genotype information is available and (b) predict mutations for family members given the probands’ confirmed genotypes. The classic and Chompret criteria are shown for comparison. Sample sizes: (a) n(mutation carriers) = 137, n(wildtypes) = 42 and (b) n(mutation carriers) = 30, n(wildtypes) = 39.

Given the confirmed genotypes of the probands, the MPC model achieved a slightly better AUC of 0.81 (95% CI, 0.70 – 0.91) (**Figure 2b**). The calibration performance of both models improved noticeably, with O/E ratios of 1.10 (95% CI, 0.80 – 1.39) and 0.96 (95% CI, 0.70 – 1.21) for the CS and MPC models, respectively. A previous validation study^36^ achieved AUCs near 0.85 when using the MPC model to predict *TP53* mutations on different research cohorts. Thus the predictive performance on clinical data is indeed lower than research data, but still at a reasonable level. With a decision threshold of 0.28, we achieved a TPR of 0.97 and a FPR of 0.51. Since the Chompret criteria are based solely on the cancer history of a family, the additional knowledge of the probands’ genotypes did not contribute toward performance. On the contrary, in this follow-up analysis, we did not make risk predictions for the probands, who often displayed strong indication for LFS, thus leading to a lower TPR of 0.23 for the Chompret criteria. These results highlight a strong advantage of our models over the standard criteria when utilizing the available information.

### Validation of cancer risk prediction

When discriminating between individuals with and without cancer, the MPC model achieved a slightly better performance compared to the KM method (AUC 0.74 vs 0.72, **Figure 3a**). When predicting SPC versus MPC, it achieved an AUC of 0.72 (95% CI, 0.61 – 0.83, **Figure 3a**). This validation included validation subjects with unknown genotypes. In practice, given the large difference in risks between the two genotype groups, it would be much more accurate to communicate the risk predictions after the patients have had confirmed testing results. Thus, we performed a similar validation, which, in addition to individuals with confirmed genotypes, included only those with *TP53* mutation probabilities that were either greater than 0.1 (inferred mutation carriers) or smaller than 0.001 (inferred wildtypes). **Figure 3b** shows an improvement in performance across all models, with the MPC model still outperforming the KM method (AUC 0.81 vs 0.77). This performance was comparable to a previous validation study^13^, which showed an AUC of 0.73 when predicting cancer versus no cancer and an AUC of 0.77 when predicting SPC versus MPC on a research cohort.

**Figure 3:**
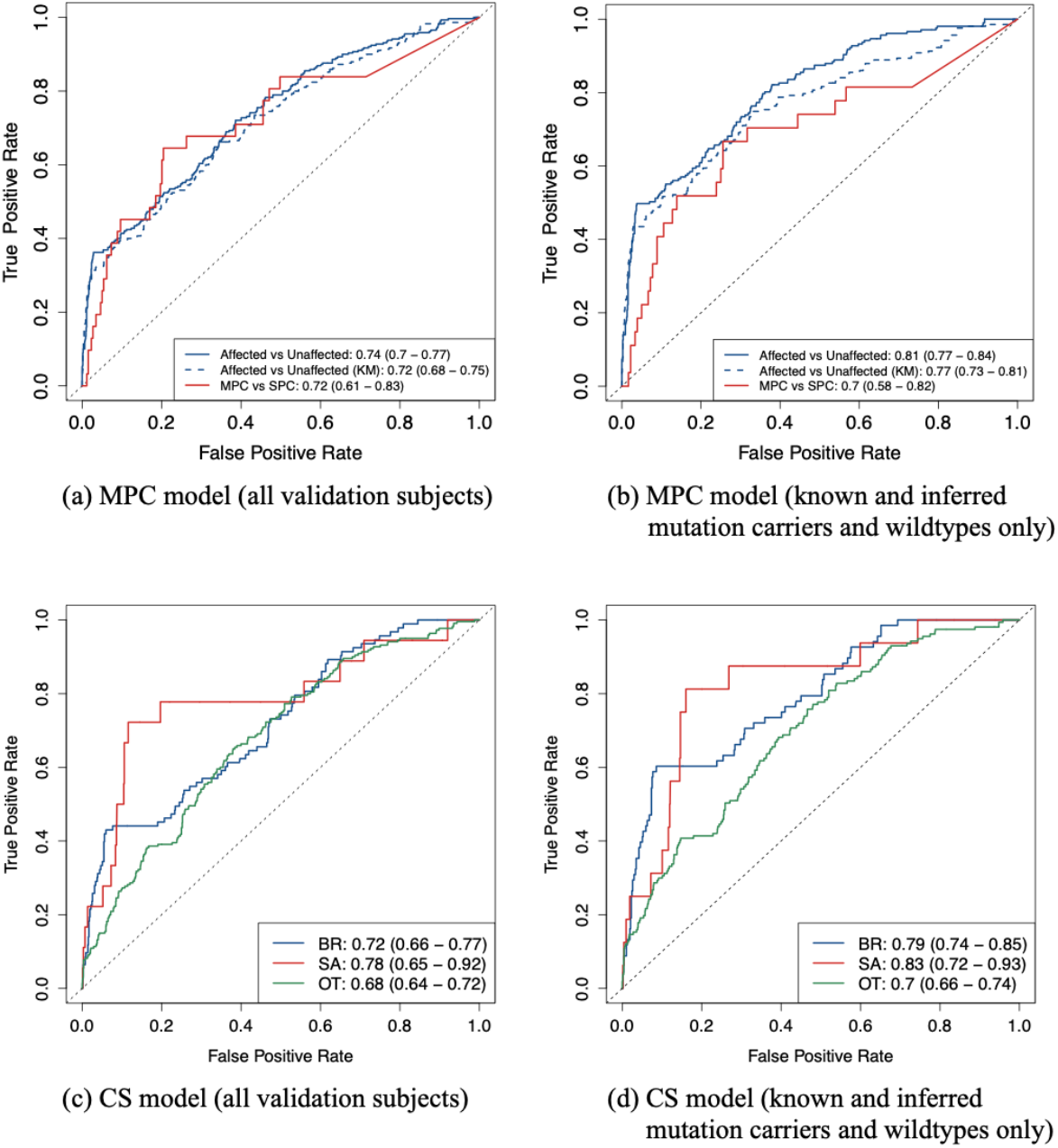
ROC curves, and the 95% bootstrapped confidence intervals of the AUCs, for predictive performance of the CS and MPC models on the CCG cohort under two scenarios: (a), (c) all validation subjects are included, and (b), (d) only known and inferred mutation carriers and wildtypes are included. For comparison, the KM method is used to predict at least one cancer versus no cancer. Sample sizes in scenario (a), (c): n(unaffected) = 1264, n(SPC) = 259, n(MPC) = 31, n(BR) = 94, n(SA) = 18, n(OT) = 220, n(D) = 497, n(A) = 879. Sample sizes in scenario (b), (d): n(unaffected) = 907, n(SPC) = 180, n(MPC) = 27, n(BR) = 69, n(SA) = 16, n(OT) = 157, n(D) = 379, n(A) = 617. BR: breast cancer, SA: sarcoma, OT: all other cancer types combined, D: death, A: alive.

The CS model achieved AUCs of 0.72, 0.78 and 0.68 for separately predicting breast cancer, sarcoma and other cancer types versus all other outcomes (**Figure 3c**). These AUCs noticeably improved to 0.79, 0.83 and 0.70, respectively, when we included only inferred mutation carriers and wildtypes in addition to genetically confirmed individuals (**Figure 3d**). Compared to validation on research cohorts^12^, we obtained a higher AUC for sarcoma, but lower AUCs for breast cancer and all other cancer types combined. Sarcoma, however, was strongly underrepresented among the validation subjects as shown in **Figure 3c** and **Figure 3d**, hence a larger sample size would be needed to make a meaningful comparison.

The calibration performances of both models were reasonably close to 1 (see **Table 3**). We observed that the O/E ratios also moved slightly towards 1 when we excluded individuals with no genetic testing results and for whom the *TP53* mutation probabilities were between 0.001 and 0.1, suggesting a potentially improved performance.

**Table 3:**
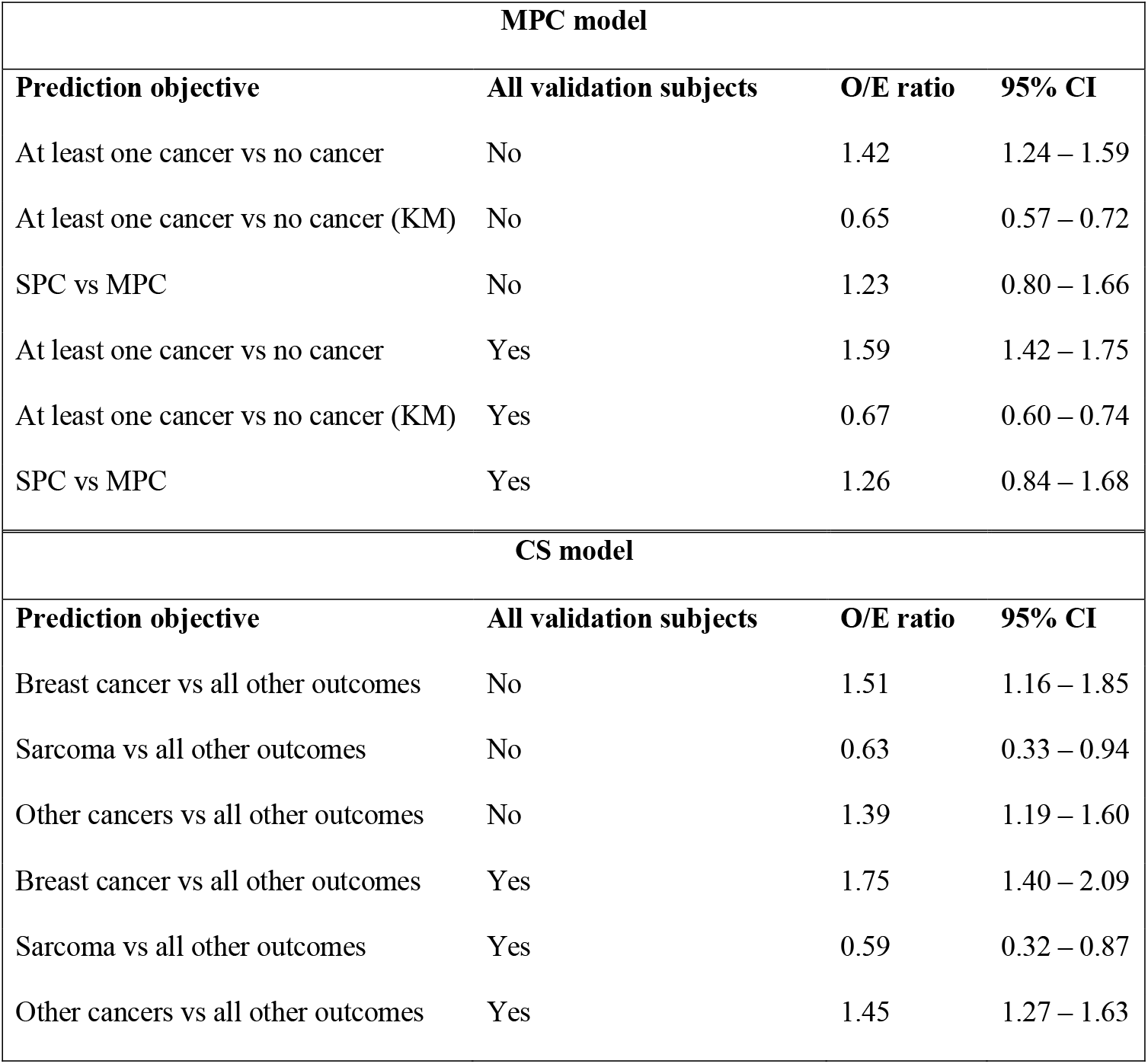
O/E ratios, along with the 95% confidence intervals, for various prediction objectives of the CS and MPC models under two scenarios: (1) all validation subjects are included (No), and (2) only known and inferred mutation carriers and wildtypes are included (Yes).

## Discussion

In this paper, we successfully conducted a unique validation of our LFS risk prediction models on a clinical counseling-based (CCB) patient cohort collected at MDACC. These models had previously been trained and validated on research protocol-based (RPB) datasets^10–13^. Our study was carefully designed to mimic scenarios that GCs encounter in real clinical settings, with 20-45% missing data, hence, to our knowledge, was the first validation study of its kind. We found that our CS and MPC models demonstrated excellent discrimination and good calibration performances when predicting deleterious germline *TP53* mutations. As expected, the performance was slightly lower than the validation results obtained using RPB cohorts^36^, most likely due to the lack of important data such as ages at last contact and ages at cancer diagnoses. For predictions of cancer risks, both models displayed performance that was comparable to previous validation studies on RPB cohorts^12,13^ in most aspects.

The strong performance of our models on the CCB cohort has important implications. It provides evidence that our research-based penetrance estimates can be accurately applied to clinical datasets that are routinely collected in counseling sessions. Our results further suggest that our models can serve as an alternative, or at least a complement, to the Chompret criteria, which are currently utilized by GCs for counseling. Finally, GCs can use our models to provide more tailored discussions based on the personalized cancer risks of their patients, rather than providing general cancer risk statistics of all LFS patients. The good calibration performance indicates that the numerical output (between 0 and 1) has a probabilistic meaning attached to it. This is important if we want to disseminate the models into clinics, since a meaningful output can aid communications between healthcare providers and patients, which have always been a challenge for rare diseases like LFS^6^.

Not only do our validation results advocate for the use of our models in clinical settings as discussed above, but they also have implications regarding clinical applications of risk prediction models in general. Given the discernible decrease in performance as we move from RPB to CCB, it is important for the research community to be aware of the differences between the two categories and, accordingly, dedicate new studies to true CCB datasets as compared to RPB datasets ^18–23^ to more accurately evaluate the real-world performance of risk prediction models. We believe that successful clinical validation studies represent a necessary step to break the barrier between model development and clinical utility. In order to bring LFSPRO closer to clinics, we aim to further perform a prospective evaluation to draw an even precise picture of how risk prediction can transform clinical practice. Lastly, the negative effects of missing data on the predictive performance highlight an important question in practice, that is whether the healthcare providers and the patients can work together to improve data collection efficiency under the time constraints in clinical sessions.

## Data Availability

All data in the present study are available contingent upon appropriate institutional data transfer agreement.

## Acknowledgements

The authors thank Dr. Gang Peng, Dr. Seung Jun Shin and Jingxiao Chen for their contributions to LFSPRO and LFSPROShiny.

## Support

Cancer Prevention and Research Institute of Texas [RP200383], National Institutes of Health [R01CA239342, P30 CA016672].

## Data sharing statement

The latest version of LFSPRO is publicly available on GitHub (https://github.com/wwylab/LFSPRO). The LFSPROShiny application is open-source on GitHub (https://github.com/wwylab/LFSPRO-ShinyApp), and is hosted live on Shinyapps.io (https://namhnguyen.shinyapps.io/lfspro-shinyapp-master/).

## Notes

Our research is supported by the Cancer Prevention and Research Institute of Texas [RP200383] and the National Institutes of Health [R01CA239342, P30 CA016672].

### Competing Interest Statement

The authors have declared no competing interest.

### Funding Statement

This study was funded by the Cancer Prevention and Research Institute of Texas [RP200383] and the National Institutes of Health [R01CA239342, P30 CA016672].

### Author Declarations

Office of Human Subject Protection/IRB of The University of Texas MD Anderson Cancer Center gave ethical approval for this work.

