## Supplementary materials for "Validating risk prediction models for multiple primaries and competing cancer outcomes in families with Li-Fraumeni syndrome using clinically ascertained data at a single institute"

### A. The training dataset

To estimate the model parameters of the CS[1] and MPC[2] models, we used a collection of 189 families that were recruited through at-risk patients at MDACC from 1944 to 1982[3–5]. These patients, called probands, were diagnosed with pediatric sarcoma before age 16 with at least three years of survival following cancer diagnoses. This dataset is ideal for model training because it was not collected specifically based on LFS criteria, hence not biased toward the classic LFS pedigree structure. Once the eligible probands had been identified through a retrospective chart review, additional phone interviews and annual follow-ups were conducted with the probands and their blood relatives to collect information on their family history, including but not limited to gender, vital status, date of birth, date of death if deceased, type of tumor and age at cancer diagnosis for all known family members. On average, three family members were contacted for each family to complete data collection. The provided information was thoroughly checked by study researchers before entering into the database: all reported deaths and cancers were confirmed with death certificates and medical records, and only invasive cancers, confirmed by either records or validated through multiple family members, were included in the finalized dataset. Mutation carrier status was defined by PCR screening of exons 2–11 of the *TP53* gene from peripheral blood cell samples of probands and additional family members that consented into the study.

### B. The CS model

In this section, we provide some important mathematical details of the CS model[1]. Let $i \in \{1, \dots, I\}$ denote the families, and $j \in \{1, \dots, n_i\}$ denote the individuals in the $i$-th family, where $n_i$ is the total number of family members. In addition, we denote by $k \in \{1, \dots, K\}$ the cancer types. We model the hazard function of the $k$-th cancer type using frailty modeling[6] as follows

$$\lambda_k\left(t \middle| \boldsymbol{X}, \xi_{i,k}\right) = \xi_{i,k}\lambda_{0,k}(t)\exp(\boldsymbol{\beta}_k^T\boldsymbol{X}), \qquad k = 1, \dots, K$$

Here $\boldsymbol{X}$ is the vector of patient-specific covariates. The choice of $\boldsymbol{X}$ is flexible depending on the research questions and the available datasets. For our application, we set $\boldsymbol{X} = \{G, S, G \times S\}^T$, where $G$ denotes the germline *TP53* mutation status (1 for deleterious mutation and 0 for wildtype) and $S$ denotes the gender (1 for male and 2 for female). We model the frailty term of the $i$-th family, $\xi_{i,k}$, via a gamma distribution that has the same shape and scale parameters: $\xi_{1,k}, \ldots, \xi_{I,k} \sim Gamma(v_k, v_k)$ independently. The importance of $\xi_{i,k}$ in accounting for within-family correlations has been validated in a previous simulation study[1]. The cumulative baseline hazard, $\Lambda_{0,k}(t) = \int_0^t \lambda_{0,k}(u)\, du$, is modeled via Bernstein polynomials, which are often used to approximate functions with constraints such as monotonicity[7]. Denoting by $M$ the degree of a Bernstein polynomial, it can be shown that[8]

$$\lambda_{0,k}(t) \approx \sum_{m=1}^{M} \gamma_{m,k} f_{B(m,M-m+1)}(t)$$

where $f_{B(m,M-m+1)}(t)$ denotes a Beta density with parameters $m$ and $M - m + 1$. Typically, we choose $M$ to be a small integer (e.g., $M = 5$) . It has been shown that Bernstein polynomials produced better performance than other choices of baseline hazard such as exponential, Weibull and piecewise functions[1].

We denote by $\boldsymbol{\theta} = \{\boldsymbol{\beta}, \boldsymbol{\gamma}, \boldsymbol{v}\}$ the vector of all model parameters, where $\boldsymbol{\beta} = \{\boldsymbol{\beta}_k : k = 1, \ldots, K\}$, $\boldsymbol{v} = \{v_k : k = 1, \ldots, K\}$ and $\boldsymbol{\gamma} = \{\gamma_{m,k} : m = 1, \ldots, M;\ k = 1, \ldots, K\}$. In addition, we write $\boldsymbol{\xi}_i = \{\xi_{i,k} : k = 1, \ldots, K\}$. For the $j$-th individual in the $i$-th family, we observe cancer history $\boldsymbol{h}_{ij} = \{t_{ij}, z_{ij}\}$, where $t_{ij}$ and $z_{ij}$ indicate the age at diagnosis and cancer type of the first primary cancer, and covariates $\boldsymbol{x}_{ij} = \{g_{ij}, s_{ij}, g_{ij} \times s_{ij}\}^T$, where $g_{ij}$ and $s_{ij}$ denote the *TP53* mutation status and sex, respectively. For individuals with no cancer occurrence, $t_{ij}$ is the censoring time, and $z_{ij} = 0$. Up to a constant of proportionality, the individual likelihood contribution is given by

$$P[\boldsymbol{h}_{ij}|\boldsymbol{x}_{ij},\boldsymbol{\theta},\boldsymbol{\xi}_i] \propto \prod_{k=1}^{K}\{\lambda_k(t_{ij}|\boldsymbol{x}_{ij},\boldsymbol{\theta},\boldsymbol{\xi}_i)\}^{\Delta_{ijk}} \times \exp\{-\Lambda_k(t_{ij}|\boldsymbol{x}_{ij},\boldsymbol{\theta},\boldsymbol{\xi}_i)\}$$

where $\Delta_{ijk} = 1$ if $\mathrm{d}_{ij} = k$ and 0 otherwise, and $\Lambda_k(t|\boldsymbol{x},\boldsymbol{\theta},\boldsymbol{\xi}_i) = \int_0^t \lambda_k(u|\boldsymbol{x},\boldsymbol{\theta},\boldsymbol{\xi}_i)\, du$ is the cancer-specific cumulative hazard. Computation of the likelihood contribution of the $i$-th family is not straight-forward, and will be discussed in **Section D**.

While the model can accommodate any number of cancer types, we restrict our attention to sarcoma, including soft tissues and osteosarcoma ($k$ = 1), breast cancer ($k$ = 2), and all other cancer types combined ($k$ = 3). We also consider mortality ($k$ = 4) as another source of competing risk, whose hazard function, $\lambda_4(\mathrm{t})$, is modeled identically. Since there are no male patients with breast cancer in the Pediatric Sarcoma cohort, we do not include gender as a covariate in the modeling of $\lambda_2(\mathrm{t})$.

### C. The MPC model

In this section, we outline the essential part of the model. For a full description of the MPC model, we refer the readers to Shin et al (2020)[2]. Let $L$ be the number of primary cancers. In statistics, MPC can be regarded as recurrent events[9]. Thus, we model cancer occurrences in patients using a non-homogenous Poisson process with intensity

$$\lambda(t|\boldsymbol{X}(t),\xi_i) = \xi_i\lambda_0(t)\exp\left(\boldsymbol{\beta}^T\boldsymbol{X}(t)\right).$$

The non-homogeneity of the Poisson process aligns with the fact that cancer risks vary significantly with age[10]. Although the choice of covariates is flexible, we use covariate vector

$\boldsymbol{X}(t) = \{G, S, G \times S, D(t), G \times D(t)\}^T$ in our application, where $G$ and S denote the genotype and gender respectively as before, and $D(t)$ is a periodically fixed indicator variable that indicates whether a patient has developed a primary cancer before time $t$. The purpose of $D(t)$ is to allow for the dependence of the subsequent primary cancers on the characteristics of the first one, which has been widely observed in many studies[11–13]. Following the CS model, we assume $\xi_1, \dots, \xi_I \sim Gamma(\nu, \nu)$ independently, and the baseline intensity is approximated as follows[8]

$$\lambda_0(t) \approx \sum_{m=1}^{M} \gamma_m f_{B(m, M-m+1)}(t),$$

where $f_{B(m,M-m+1)}(t)$ denotes a Beta density with parameters $m$ and $M - m + 1$.

We denote by $\boldsymbol{\theta} = \{\boldsymbol{\beta}, \boldsymbol{\gamma}, \nu\}$ the vector of all model parameters, where $\boldsymbol{\gamma} = \{\gamma_m : m = 1, \dots, M\}$. For the $j$-th individual in the $i$-th family, let $L_{ij}$ be the number of cancer occurrences. We observe cancer history $\boldsymbol{h}_{ij} = \{\boldsymbol{t}_{ij}, \boldsymbol{z}_{ij}, c_{ij}\}$, where $\boldsymbol{t}_{ij} = \{t_{ij,l} : l = 1, \dots, L_{ij}\}^T$ and $\boldsymbol{z}_{ij} = \{z_{ij,l} : l = 1, \dots, L_{ij}\}^T$ are $L_{ij}$-dimensional vectors that contain the ages at diagnosis and cancer types of the primary cancers, and $c_{ij}$ is the censoring time. For individuals with at least one primary cancer, we define $d_{ij}(t) = 1$ if $t > t_{ij,1}$ and 0 otherwise. For individuals with no cancer occurrences, $d_{ij}(t) = 1$ for all $t \in [0, c_{ij}]$. Then, the vector of covariates is given by $\boldsymbol{x}_{ij}(t) = \{g_{ij}, s_{ij}, g_{ij} \times s_{ij}, d_{ij}(t), g_{ij} \times d_{ij}(t)\}^T$, where $g_{ij}$ and $s_{ij}$ denote the *TP53* mutation status and sex, respectively, as before. For convenience, we let $t_{ij,0} = 0$. Up to a constant of proportionality, the individual likelihood contribution is given by

$$P\left[\boldsymbol{h}_{ij} \middle| \boldsymbol{x}_{ij}(t), \boldsymbol{\theta}, \xi_i\right] \propto \left\{ \prod_{l=1}^{L_{ij}} \lambda\left(t_{ij,l} \middle| \boldsymbol{x}_{ij}(t_{ij,l-1}), \boldsymbol{\theta}, \xi_i\right) \right\} \times$$

$$exp\left\{-\sum_{l=1}^{L_{ij}}\Lambda\big(t_{ij,l}\big|\boldsymbol{x}_{ij}(t_{ij,l-1}),\boldsymbol{\theta},\xi_i\big)\right\}\times\exp\left\{-\Lambda\left(c_{ij}\middle|\boldsymbol{x}_{ij}(t_{ij,L_{ij}}),\boldsymbol{\theta},\xi_i\right)\right\}$$

where $\Lambda\big(t\big|\boldsymbol{x}_{ij}(u),\boldsymbol{\theta},\xi_i\big)=\int_u^t\lambda\big(v\big|\boldsymbol{x}_{ij}(u),\boldsymbol{\theta},\xi_i\big)\,dv$.

While our model can work with any number of primary cancers, we only model up to the second primary (i.e., $L=2$) due to the limited occurrences of a third primary cancer and beyond.

### D. Model estimation

The process of estimating the model parameters is likelihood-based, and similar, albeit not identical, for the CS and MPC models. In previous sections, we have computed the individual likelihood contribution of a family member. Computation of the family-wise likelihood, however, is not straightforward since most family members do not undergo genetic testing and are thus related through missing genotypes. In this case, knowledge of a family member's genotype is informative about other family members, hence we cannot consider the individuals as being independent. For this reason, the family-wise likelihood cannot be simply computed as the product of the individual likelihood contributions from all the family members. We consider a family with $n$ family members. Let $\boldsymbol{H}=\{H_1,\dots,H_n\}$ be the cancer history and $\boldsymbol{G}=\{G_1,\dots,G_n\}$ be the family's genotype data. We partition $\boldsymbol{G}=\boldsymbol{G}_{obs}\cup\boldsymbol{G}_{obs}^c$, where $\boldsymbol{G}_{obs}$ denotes the observed part of the genotype data (i.e., $\boldsymbol{G}_{obs}^c$ denotes the missing part). We use the Elston-Stewart peeling algorithm[14] to compute the family-wise likelihood $P[\boldsymbol{H}|\boldsymbol{G}_{obs}]$ recursively in a way that accounts for the pedigree structure (**Supplementary Figure 1**). Given a pivot member, the algorithm splits the pedigree tree into two disjoint groups: (i) the anterior, which consists of family members that are related to the pivot through the children and spouse, and (ii) the posterior, which consists of those that are related to the pivot through the parents. These groups are connected only through the pivot, hence conditionally independent given the pivot's genotype. If the pivot member has unknown genotype, it can be shown that

$$P[\boldsymbol{H}|\boldsymbol{G}_{obs}] = \sum_{G_p \in \{0,1\}} P\left[\boldsymbol{H}_p^-|G_p, \boldsymbol{G}_{obs}\right] P\left[H_p|G_p\right] P\left[\boldsymbol{H}_p^+|G_p, \boldsymbol{G}_{obs}\right]$$

where $G_p$ denotes the genotype of the pivot, $\boldsymbol{H}_p^-$ denotes the cancer history of the anterior, and $\boldsymbol{H}_p^+$ denotes the cancer history of the posterior. If the pivot member has confirmed genotype, we can simply remove the summation in the expression above. The term $P\left[H_p|G_p\right]$ is the individual likelihood contribution of the pivot member, which is available in closed form for both the CS and MPC models. To compute the anterior and posterior likelihoods, the algorithm follows the same steps (i.e., randomly pick a pivot member within each group, and split the group into two smaller subgroups). Eventually, the computation boils down to individual likelihoods, which can be readily computed from the proposed models.

**[Supplementary Figure 1]**

Another major challenge is to correct for ascertainment bias, which arises because we sample the patients from high-risk clinics to ensure good sample size. To overcome this problem, we adopt the ascertainment-corrected joint (ACJ) likelihood approach[15]. Let $\mathcal{A}$ be the ascertainment indicator (i.e., $\mathcal{A} = 1$ if a family is ascertained and 0 otherwise). Under some reasonable assumptions, it follows that

$$P[\boldsymbol{H}, \boldsymbol{G}_{obs}|\mathcal{A} = 1] \propto \frac{P[\boldsymbol{H}|\boldsymbol{G}_{obs}]}{P[\mathcal{A} = 1]}$$

Thus, given the family-wise likelihood from the peeling algorithm, we inversely scale it by the ascertainment probability. Calculations of the ascertainment probability depend on the training dataset. Each family in the Pediatric Sarcoma dataset starts with a proband, who was diagnosed with pediatric sarcoma at MD Anderson Cancer Center. Due to suspicion of a hereditary cancer syndrome that ran through the family, data collection was then extended to other family members through phone interviews and extended follow-ups. Hence, in both models, we calculate the ascertainment probability as the likelihood contribution of the proband.

Once the ACJ likelihoods has been computed for all the families, the overall likelihood of the dataset is simply given by their product since the families can be considered independent. We use the Metropolis-Hasting-within-Gibbs algorithm to generate 100,000 posterior samples, with the first 10,000 discarded as burn-in. The model parameters are then estimated as the means or medians of the remaining posterior samples.

### E. Computation of carrier probability

Given a counselee with unknown genotype $G_0$ and history $\boldsymbol{H} = \{H_1, \dots, H_n\}$ of the $n$ family members, our goal is to estimate $P[G_0|\boldsymbol{H}]$. To do this, we follow our previous study[16] and set the prevalence of pathogenic *TP53* mutations in the general population to be 0.0006. Assuming the Hardy-Weinberg equilibrium, it follows that the prevalence of wildtype ($G_0 = 0$), heterozygous mutation ($G_0 = 1$) and homozygous mutation ($G_0 = 2$) are 0.9988, 0.0005996 and 3.6e-07, respectively. Our models compute the posterior probabilities $P[G_0 = g|\boldsymbol{H}]$, $g \in \{0,1,2\}$, via the Bayes rule based on Mendelian transmission as follows

$$P[G_0|\boldsymbol{H}] = \frac{P[G_0]P[\boldsymbol{H}|G_0]}{\sum_{G_0} P[G_0]P[\boldsymbol{H}|G_0]}$$

where

$$P[\boldsymbol{H}|G_0] = \sum_{G_1,\dots,G_n} P[\boldsymbol{H}|G_0, G_1, \dots, G_n] P[G_1, \dots, G_n|G_0]$$

$$= \sum_{G_1,\dots,G_n} \left[ \prod_{j=0}^{n} P[H_j|G_j] \right] P[G_1, \dots, G_n|G_0]$$

We can assume that the family members are conditionally independent given all the confirmed genotypes, hence the family-wise likelihood $P[\boldsymbol{H}|G_0, G_1, \dots, G_n]$, factorizes into a product of the individual likelihoods $P[H_j|G_j]$, $j \in \{1, \dots, n\}$, whose analytical expressions in the CS and MPC models have been given in **Section B** and **Section C**, respectively. Assuming Mendelian transmission, computation of $P[G_1, \dots, G_n|G_0]$ can be very complex for large families. To overcome this computational issue, we employ the peeling algorithm[14], which has been discussed in **Section D**, to calculate $P[\boldsymbol{H}|G_0]$ recursively in an efficient way.

### F. Parameter estimates of the CS and MPC models

Following the procedure described in Section 3, we fitted the CS and MPC models to the Pediatric Sarcoma dataset. **Supplementary Table 1** and **Supplementary Table 2** show estimates of the regression coefficients.

**[Supplementary Table 1]**

**[Supplementary Table 2]**

### G. The prospective cohort at MD Anderson Cancer Center

This is a research cohort that was collected at MD Anderson Cancer Center (MDACC) from year 2000 to 2015. It consists of prospectively followed families, each of which was recruited through a family member that was considered to be at high risks of deleterious germline *TP53* mutations based on the classic LFS criteria[17] or the Chompret criteria[18–20]. These family members, called the probands, were identified by trained personnel in the Department of Genetics via one of the following channels: (1) surgery schedules, (2) ClinicStation, (3) patient census, (4) patient clinics, (5) patient-study areas and referrals from inside or outside MDACC, and (6) self-referrals. Upon approval by the principal investigator, the probands were invited to participate in the study. The data collection procedure was expanded to the probands' family members in a similar way to the Pediatric Sarcoma dataset, resulting in an extensive cohort of 11,186 individuals spanning across 429 families. We will refer to this dataset as the MDACC prospective cohort. Summary of the dataset is displayed in **Supplementary Table 3**.

**[Supplementary Table 3]**

### H. Comparison of MDACC prospective and CCG datasets

The MDACC prospective cohort was previously used to validate the risk predictions of the CS and MPC models[1,2]. While the validation studies led to successful results, there are fundamental differences between research and clinical cohorts. **Supplementary Table 4** highlights the differences between the two datasets. As in the case of the Pediatric Sarcoma dataset, we notice that the MDACC prospective cohort consists of families that have over 100 family members due to the rigorous data collection protocols (e.g., phone interviews, regular follow-ups). Furthermore, it is clear that the MDACC prospective cohort has complete ages at last contact and ages at cancer diagnoses across all family members.

**[Supplementary Table 4]**

### Supplementary tables/figures

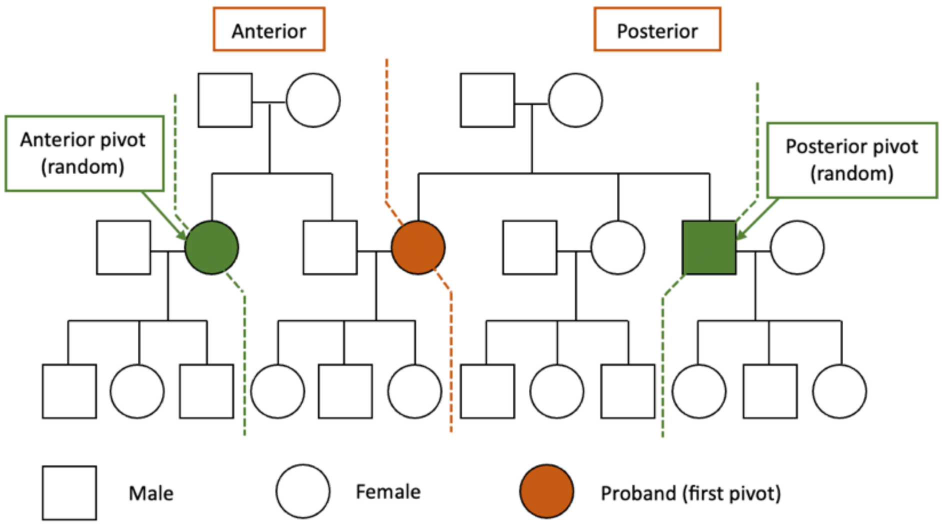


*Supplementary Figure 1: Illustration of the peeling algorithm, which uses recursion to compute the family-wise likelihood while accounting for the familial structure.*

| Cancer type | Parameter | Mean | Standard deviation | 2.5% | 97.5% |
|---|---|---|---|---|---|
| Breast | $\beta_G$ | 3.560 | 0.561 | 2.541 | 4.544 |
| Sarcoma | $\beta_G$ | 2.464 | 0.895 | 0.675 | 4.182 |
| | $\beta_S$ | -3.677 | 1.077 | -6.176 | -1.902 |
| | $\beta_{G\times S}$ | 0.971 | 0.548 | -0.110 | 2.040 |
| Other cancers | $\beta_G$ | 1.576 | 0.769 | 0.072 | 3.072 |
| | $\beta_S$ | -0.993 | 0.186 | -1.366 | -0.647 |
| | $\beta_{G\times S}$ | 0.559 | 0.574 | -0.620 | 1.628 |

*Supplementary Table 1: Mean estimates and 95% credible interval of the regression coefficients $\boldsymbol{\beta}_k$ in the CS model*

| Parameter | Median | Standard deviation | 2.5% | 97.5% |
|---|---|---|---|---|
| $\beta_G$ | 3.516 | 0.256 | 3.068 | 3.953 |
| $\beta_S$ | 0.027 | 0.115 | -0.189 | 0.232 |
| $\beta_{G\times S}$ | -0.332 | 0.246 | -0.809 | 0.139 |
| $\beta_{D(t)}$ | -0.380 | 0.363 | -1.152 | 0.259 |
| $\beta_{G\times D(t)}$ | 0.716 | 0.429 | -0.070 | 1.601 |

*Supplementary Table 2: Median estimates and 95% credible interval of the regression coefficients $\boldsymbol{\beta}$ in the MPC model*

| | MDACC prospective | | | |
|---|---|---|---|---|
| | Wildtype | Mutation | Unknown | Total |
| **Male** | | | | |
| Healthy | 119 | 41 | 4,448 | 4,601 |
| SPC | 58 | 50 | 747 | 855 |
| MPC | 33 | 39 | 60 | 132 |
| Subtotal | 210 | 130 | 5,255 | 5,595 |
| **Female** | | | | |
| Healthy | 144 | 32 | 4,116 | 4,292 |
| SPC | 113 | 76 | 798 | 987 |
| MPC | 114 | 97 | 101 | 312 |
| Subtotal | 371 | 205 | 5,015 | 5,591 |
| **Total** | 581 | 335 | 10,270 | 11,186 |

*Supplementary Table 3: Categorization of family members in the MDACC prospective dataset by gender, number of primary cancers and mutation status. SPC = single primary cancer, MPC = multiple primary cancer*

| | MDACC prospective | CCG |
|---|---|---|
| **Number of families** | | |
| **All family members** | | |
| Complete data | 429 (100%) | 10 (8%) |
| Missing ages at last contact only | 0 (0%) | 46 (37%) |
| Missing ages at cancer diagnosis only | 0 (0%) | 0 (0%) |
| Missing both ages at last contact and ages at cancer diagnosis | 0 (0%) | 68 (55%) |
| **Total** | **429** | **124** |
| **Chi-squared test** | **P < 0.001** | |
| **First-degree relatives and spouse only** | | |
| Complete data | 429 (100%) | 68 (55%) |
| Missing ages at last contact only | 0 (0%) | 41 (33%) |
| Missing ages at cancer diagnosis only | 0 (0%) | 10 (8%) |
| Missing both ages at last contact and ages at cancer diagnosis | 0 (0%) | 5 (4%) |
| **Total** | **429** | **124** |
| **Chi-squared test** | **P < 0.001** | |
| **Number of individuals** | | |
| **All family members** | | |
| Complete data | 11,186 (100%) | 1,748 (53%) |
| Missing ages at last contact only | 0 (0%) | 1,339 (41%) |

| | | |
|---|---|---|
| Missing ages at cancer diagnosis only | 0 (0%) | 138 (4%) |
| Missing both ages at last contact and ages at cancer diagnosis | 0 (0%) | 72 (2%) |
| **Total** | **11,186** | **3,297** |
| **Chi-squared test** | **P < 0.001** | |
| **First-degree relatives and spouse only** | | |
| Complete data | 2,654 (100%) | 487 (79%) |
| Missing ages at last contact only | 0 (0%) | 105 (17%) |
| Missing ages at cancer diagnosis only | 0 (0%) | 19 (3%) |
| Missing both ages at last contact and ages at cancer diagnosis | 0 (0%) | 2 (< 1%) |
| **Total** | **2,654** | **613** |
| **Chi-squared test** | **P < 0.001** | |
| **Number of individuals per family** | | |
| Min | 3.00 | 1.00 |
| 5% percentile | 6.00 | 1.00 |
| 10% percentile | 9.80 | 4.00 |
| 25% percentile | 14.00 | 16.00 |
| Median | 22.00 | 26.50 |
| Mean | 28.63 | 26.59 |
| 75% percentile | 34.00 | 36.00 |
| 90% percentile | 54.00 | 48.00 |
| 95% percentile | 77.60 | 53.85 |

| | | |
|---|---|---|
| Max | 151.00 | 75.00 |

*Supplementary Table 4: Comparison of a research cohort (MDACC prospective) and a clinical cohort (CCG) on the extent of missing ages at last contact and missing ages at cancer diagnoses at both family and individual levels. Summary statistics for the number of individuals per family are reported to contrast the depth of data collection procedures in research and clinical cohorts as they happen in the unit of families*
